# Emergence of porcine delta-coronavirus pathogenic infections among children in Haiti through independent zoonoses and convergent evolution

**DOI:** 10.1101/2021.03.19.21253391

**Authors:** John A. Lednicky, Massimiliano S. Tagliamonte, Sarah K. White, Maha A. Elbadry, Md. Mahbubul Alam, Caroline J. Stephenson, Tania S. Bonny, Julia C. Loeb, Taina Telisma, Sonese Chavannes, David A. Ostrov, Carla Mavian, Valerie Madsen Beau De Rochars, Marco Salemi, J. Glenn Morris

## Abstract

Coronaviruses have caused three major epidemics since 2003, including the ongoing SARS-CoV-2 pandemic. In each case, coronavirus emergence in our species has been associated with zoonotic transmissions from animal reservoirs^1,2^, underscoring how prone such pathogens are to spill over and adapt to new species. Among the four recognized genera of the family *Coronaviridae* – *Alphacoronavirus, Betacoronavirus, Deltacoronavirus, Gammacoronavirus*, – human infections reported to date have been limited to alpha- and betacoronaviruses^3^. We identify, for the first time, porcine deltacoronavirus (PDCoV) strains in plasma samples of three Haitian children with acute undifferentiated febrile illness. Genomic and evolutionary analyses reveal that human infections were the result of at least two independent zoonoses of distinct viral lineages that acquired the same mutational signature in the *nsp15* and the *spike* glycoprotein genes by convergent evolution. In particular, structural analysis predicts that one of the changes in the Spike S1 subunit, which contains the receptor-binding domain, may affect protein’s flexibility and binding to the host cell receptor. Our findings not only underscore the ability of deltacoronaviruses to adapt and potentially lead to human-to-human transmission, but also raise questions about the role of such transmissions in development of pre-existing immunity to other coronaviruses, such as SARS-CoV-2.

---

Coronaviruses are enveloped, positive-sense single-stranded RNA viruses that belong to the family *Coronaviridae*, subfamily *Coronavirinae*. They are responsible for respiratory syndromes and hepatic and gastroenteric diseases in birds and mammals^4^. Their host cell binding protein, the Spike (S) glycoprotein, binds to receptors relatively conserved across species^5,6^; this characteristic can facilitate cross-species transmission, and lead to outbreaks and endemicity in the human population, as has already happened multiple times^1,2^. Their propensity for recombination^1,5,7,8^ can also facilitate the escape from local fitness optima and broaden their species tropism^1,5,8-10^.

Porcine deltacoronavirus (PDCoV) is a member of the genus *Deltacoronavirus*, and the virus was reported for the first time in Hong Kong, China, in 2012^11^, and later in the Americas, in 2014^12^. It causes gastrointestinal symptoms in piglets, with transient viremia, which may lead to dehydration and death^13,14^. The virus infects villous epithelial cells of the entire large intestine, although the jejunum and ileum are the primary sites of infection^13,15^. Studies^6,16^ have demonstrated that PDCoV employs the host aminopeptidase N (APN) protein as an entry receptor through an interaction with it *via* domain B of its spike (S) glycoprotein^6^. Further cause of concern are reports of symptomatic infection, in experimental settings, of chickens and turkeys^17^. Human cells were also reported to be permissive to PDCoV infection ^6^. Such evidence suggests that the binding of the PDCoV S glycoprotein to an interspecies conserved site (APN protein) may facilitate direct transmission to non-reservoir species, possibly including humans.

Haiti is part of the island of Hispaniola, and one of the poorest countries in the world^18^. Efforts were made to wipe out the local pig population in the 1980s to eliminate African swine fever from the area^19^. Pigs were then reintroduced from North American populations, mainly the USA and in small part, Canada^19,20^. Pig farming in the country is at a subsistence level, and, to our knowledge, no PDCoV cases have been reported to date in pigs. Our group monitored occurrence of illness among children attending school within a school system operated by the Christianville Foundation in the Gressier region of Haiti from 2012 to 2020^21-24^. During that time period, free medical care was provided to children at the school clinic. Among children seen at the clinic, respiratory and diarrheal illnesses were most common, followed by acute undifferentiated febrile illnesses (fever with no clear localizing symptoms) that were reported by approximately 16% of clinic attendees^21^.

## Detection of human-PDCoV (Hu-PDCoV) strains in Haitian pediatric patients

This study was approved by the Institutional Review Board (IRB) at the University of Florida and the Haitian National IRB; written informed consent for sample collection was obtained from parents of participants, with assent from participants. Beginning in May 2014 plasma samples were collected from children presenting to the school clinic with acute undifferentiated febrile illness, with a total of 369 samples collected between May 2014 and December 2015. As described below, three of these children (aged 5-10 years) were subsequently found to be infected with coronavirus strains which clustered with PDCoV. Cases 1 and 3 were in patients from the main campus of the school (School A) that is attended by students from semi-urban areas, with students coming from a range of socioeconomic backgrounds. Case 2 was from a different campus (School B), which is located in the mountains approximately an hour’s drive from School A, in a completely rural area, with students from very low socioeconomic backgrounds. All three children presented with a history of fever but recovered uneventfully: Child 2 was febrile (40°C) when seen in the clinic; Child 1 and 2 reported cough and abdominal pain. Child 3, while reporting a fever, did not have acute symptoms when seen in clinic.

Nucleic acids purified as previously described^25,26^ from the plasma samples of the three children of Table 1 tested negative for alpha- and flavivirus RNAs^25,26^ by RT-PCR. To screen for viruses not detected by the RT-PCR, virus isolation was also attempted after inoculation of aliquots of the plasma onto Vero E6 cells^25,26^. Nucleic acids, purified from the cell culture media7, 14, 21, and 30 days post-inoculation (dpi) of the cells, again tested negative for alpha- and flavivirus RNAs. Moreover, they tested negative for the DNA and RNA of common human respiratory viruses using a GenMark Respiratory Panel^27^. However, subtle cytopathic effects (CPEs) were observed in Vero E6 cell monolayers starting at about 11 dpi, suggesting that a virus had been isolated. The non-specific CPEs included granulation of the cells (Fig. 1). Since none of the tests produced evidence that could be used for a preliminary identification of a viral agent, an unbiased amplification and sequencing approach^28^ was attempted for cells inoculated with plasma from sample 0081-4, which displayed more CPEs than cells inoculated with the other two plasma samples. PCR amplification yielded seven amplicons. Sequence analyses indicated that six were African green monkey sequences from the Vero E6 cells, whereas one 401 bp amplicon had 100% identity with the corresponding genome sequence of various PDCoV strains, such as CHN-SC2015 (GenBank MK355396). Therefore, RNA purified from Vero E6 culture samples was re-tested using a pancoronavirus RT-PCR test that amplifies a 668 bp-region within the RNA-dependent RNA polymerase gene that encodes the most conserved protein domain of alpha-, beta-, gamma-, and delta CoVs^29^, generating positive results. A follow-up test using RNA directly purified from plasma generated the same 668-bp amplicons, providing further indications that a PDCoV was present in the plasma and cell culture samples. In contrast, 20 other plasma samples and mock-inoculated Vero E6 cultures tested negative for PDCoV RNAs. At the time, we had no PDCoV strains in our laboratory. Following this preliminary identification, whole genome sequences for the three isolates were obtained by Sanger sequencing. The GenBank accession numbers corresponding to the sequenced genomes are given in Table 1.

**TABLE 1.** Sample data, children with PDCoV infection.

| Child case # | Collection date | School | PDCoV strain | GenBank No. | GenBank No. of closest PDCoV strain by BLAST analysis |
| --- | --- | --- | --- | --- | --- |
| 1 | Dec. 2014 | School A | Haiti/Human/0081-4/2014 | MW685622 | KY065120 (Pig/China/2016) |
| 2 | March 2015 | School B | Haiti/Human/0256-1/2015 | MW685623 | KR150443 (Pig/USA/2015) |
| 3 | April 2015 | School A | Haiti/Human/0329-4/2015 | MW685624 | KY065120 (Pig/China/2016) |

**Figure 1:**
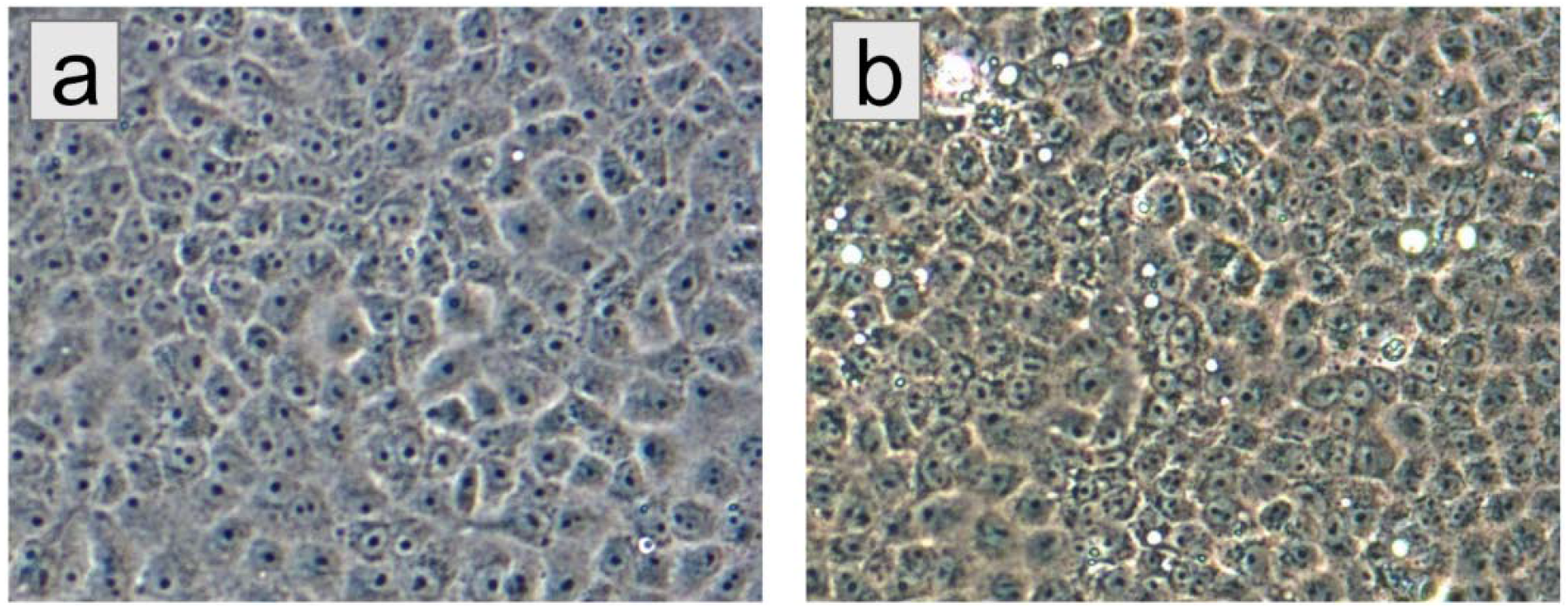
Non-specific CPEs formed by plasma from patient 0081-4 in Vero E6 cells. a) Mock-infected Vero E6 cells, 11 days port-inoculation with phosphate-buffered saline. b) Vero E6 cells 11 days post-inoculation with plasma from patient 0081-4. Original magnification at 200X

### Genomic characterization and evolution of Hu-PDCoV strains

Assessment of potential recombinants ^30,31^ in a multiple sequence alignment including currently available full genome PDCoV sequences did not detect any signal for recombination in the viral strains from human patients (Supplemental Figures S1a, S1b), and displayed robust signal (Supplemental Figure S1c) for phylogeny inference. Strains from case 1 (designated as strain 0081-4) and case 3 (strain 0329-4), identified four months apart among children attending School A (Table 1), were highly similar (99.97%) and closely related (99.8%) to a pig strain detected one year later in Tianjin, China. Case 3 (strain 0246-1), who attended School B in the mountains, was infected with a variant strain similar to the other two Hu-PDCoV strains (98.9%), but most closely related to a pig strain detected in Arkansas, USA, in 2015. Despite such a high genomic similarity, the maximum likelihood (ML) tree inferred from full genome sequences clearly shows that strains from cases 1 and 3 and the strain from case 2 belong, respectively, to two distinct and well supported monophyletic clades: the first one clustering strains from Chinese pigs, and the second one clustering strains from USA pigs (Fig. 2). In other words, both epidemiology data and evolutionary relationships indicate that the three PDCoV strains identified in the Haitian children were the result of at least two separate zoonotic transmissions that likely occurred within a similar time frame. The close evolutionary relationships of strain 0256-1 with a USA PDCoV sequence sampled in 2014-2015 is not surprising given the re-introduction of pigs from USA to Haiti after the local population was mostly wiped out following the African Swine Fever epidemic^19,20^. On the other hand, the close phylogenetic relationship between Haitian strains 0081-4 and 0329-4 and a viral sequence isolated from a Chinese pig one year later (Table 1) is less obvious. These three sequences clearly share a most recent common ancestor (MRCA) with high bootstrap support (99%). The Haitian PDCoV strains may have been transmitted to the children through two independent zoonoses from the same animal, or different animals infected with a highly similar virus, or be the result of an initial zoonosis followed by human-to-human transmission. Since samples from pigs in the areas surrounding the two schools were not available, it is impossible at this time to discern which scenario is the most likely. Regardless, the phylogeny demonstrates the circulation of two distinct PDCoV lineages in School A and School B, highlighting deltacoronaviruses’ ability to spill over successfully in the human population.

**Figure 2.**
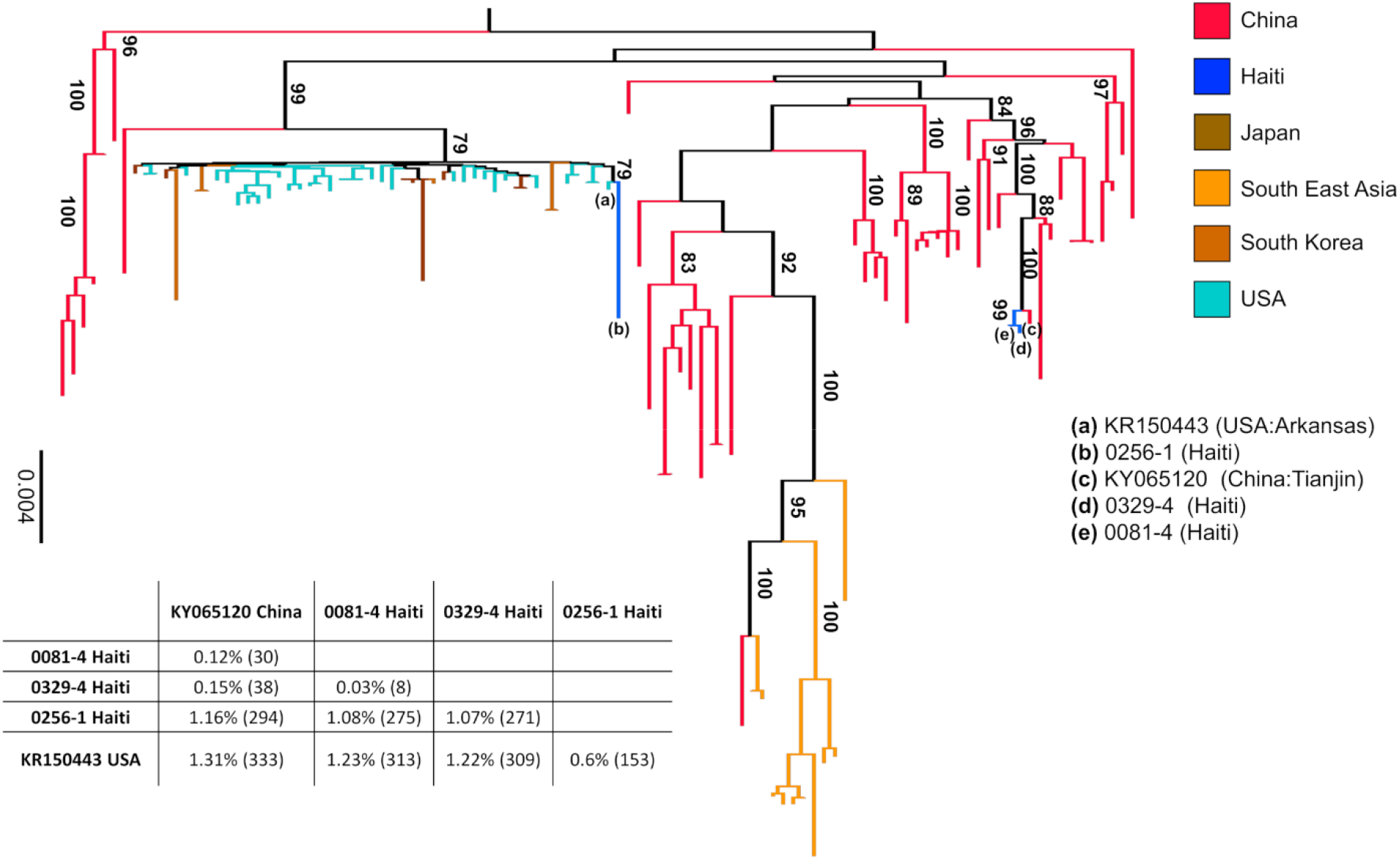
Maximum likelihood tree of PDCoV strains. The tree was inferred from 109 PDCoV full genome sequences, including four sparrow CoV genomes for outgroup rooting (accession numbers MG812375, MG812376, MG812377, and MG812378, from Chen *et al*., 2018) using the best fitting nucleotide substitution model (see Methods). For display purposes, outgroup sequences were removed from the figure. Vertical branches are scaled in number of nucleotide substitutions per site according to the bar on the left of the tree. Bootstrap values > 75% (1000 replicates) are indicated along supported branches. The table at the bottom of the tree shows nucleotide % dissimilarity (and total number of nucleotide differences in parenthesis) between Haitian strains and their closest non-Haitian relatives, which are labelled in tree by progressive letters according to the legend on the right.

Our next step was the calibration of a molecular clock to infer the time of the most recent ancestor (TMRCA) of Hu-PDCoV and their most closely related porcine strains. We tested for the presence of temporal signal in the sequence data set by calculating the linear regression between root-to-tip distances and sampling time in the ML tree^32^. After removal of sparrow outgroup sequences and the South East Asian clade outliers, the tree inferred from the remaining (n = 94) sequences showed sufficient signal to calibrate a molecular clock (see Supplemental Figure S2 for details). The topology of the Bayesian maximum clade credibility (MCC) tree obtained by enforcing a strict molecular clock confirmed the findings of the ML phylogeny (Fig. 3). Identical results were obtained by enforcing a relaxed clock model. According to the clock calibration, 0081-4 and 329-4 TMRCA dates back to October 2014, with 95% high posterior density intervals (95%HPD) essentially overlapping (October 2014 – January 2015) with the sampling dates (see Table 1) of the strains themselves. In turn, the Haitian strains diverged from their MRCA with the pig Chinese strain in July 2014 (95%HPD: April – August 2014). These results point to a recent introduction in the country before the virus infected the children and are compatible both with zoonotic events and/or limited human-to-human transmission. On the other hand, Haitian isolate 0256-1 TMRCA with USA isolate KR150443 dates back to 2011 (95% HPD: Feb-2011 - March-2012). Therefore, the strain had likely been circulating in pigs for a few years, as also suggested by its relatively long terminal branch in the ML tree (Fig. 2), before infecting the human patient. It is possible, of course, that we are missing several intermediate links along the 0256-1 branch, either from pig or other human strains, and the introduction in the country might have been more recent than suggested by the molecular clock analysis.

**Figure 3.**
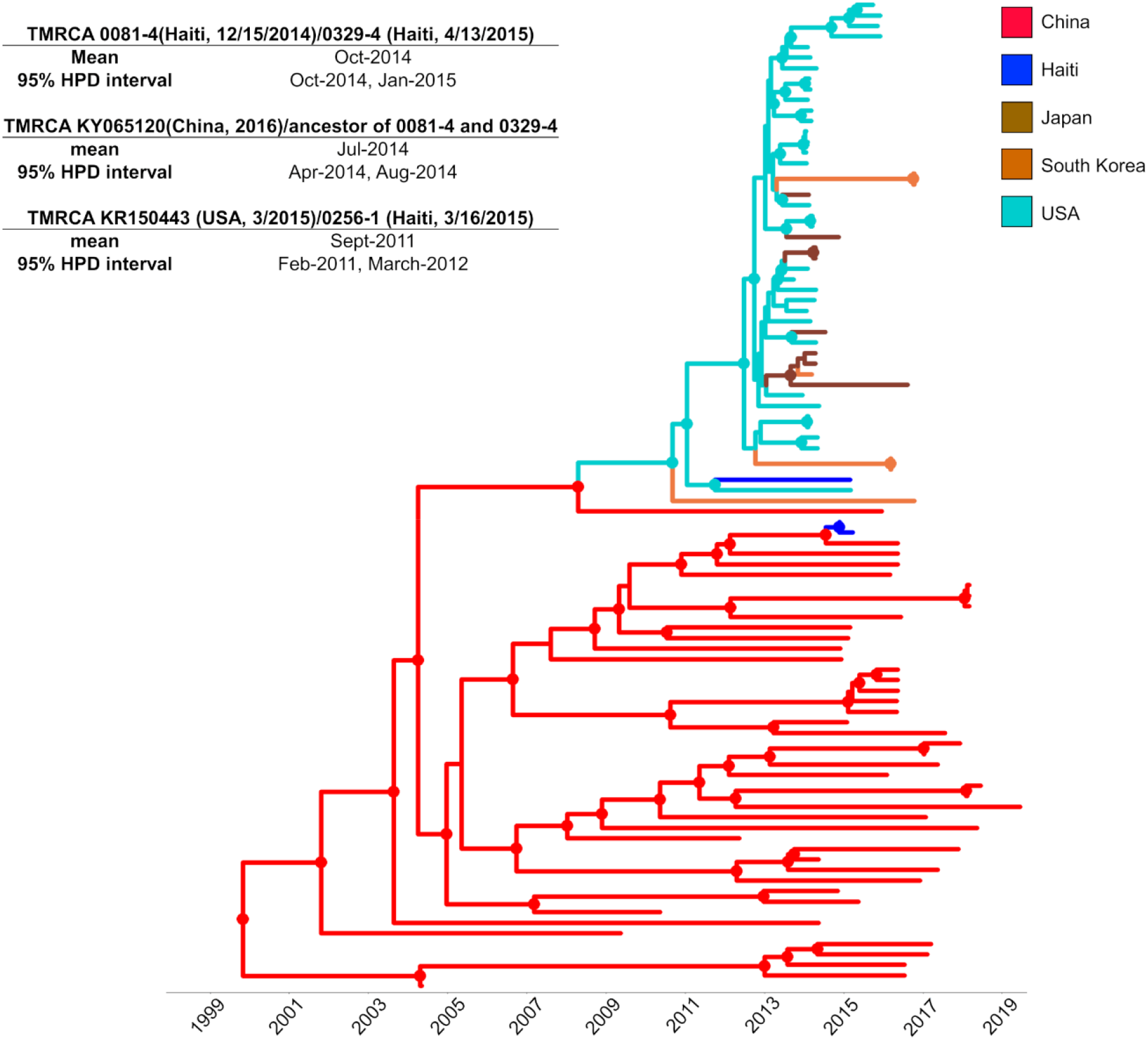
Bayesian maximum clade credibility (MCC) tree of PDCoV strains. The MCC tree was inferred from a subset of 94 full genome strains that displayed sufficient temporal signal for molecular clock calibration. Branch lengths were scalded in time, according to the bar at the bottom, by enforcing a strict molecular clock and using sampling dates to estimate PDCoV evolutionary rate. Circles at internal nodes indicate high posterior probability (PP) support >0.9. The table on the left shows the inferred time of the most recent common ancestor (TMRCA) between Haitian strains and their closest phylogenetic relative, with 95% high posterior density intervals (95%HPD).

While phylogeny inference demonstrates that the Hu-PDCoV strains isolated from Haitian children belong to independent evolutionary lineages, introduced in humans through at least two separate zoonotic transmissions, a more in-depth analysis of the genomic changes occurred along the human lineages revealed an interesting pattern. Signature pattern analysis^33^ shows that the three Haitian strains share a signature of five conserved amino acid residues in the ORF1a/b polyprotein and two in the Spike glycoprotein, unique among other PDCoV sequences from pigs currently known (Fig. 4a). The sole exception is Chinese strain KY065120, which displays the same amino acid signature and is the one most closely related to Haitian strains from School A (Fig. 2 and Table 1) and may represent a porcine strain pre-adapted for effective transmission to humans. Indeed, the convergent evolution of identical amino acid changes along distinct phylogenetic lineages is highly suggestive of an adaptive response. Mutations in the first five ORF1/ab amino acids part of the Hu-PDCoV-specific signature (Fig. 4a) are located at sites that do not correspond to solved crystal structures. The other ORF1/ab mutation maps in the nonstructural protein 15 (Nsp15): A30V (amino acid position numbered according to reference sequence JQ065043). The C-terminal domain of the Nsp15 protein possesses endoribonuclease with uridylate-specific activity^34^. While the protein is not necessary for RNA synthesis, it is necessary in coronaviruses to escape recognition of dsRNA intermediates by the host^35^. PDCoV Nsp15 inhibits the induction of IFN-β, the main intestinal antiviral cytokine, by preventing interferon regulatory factors IRF1 nuclear translocation^36^. The last two mutations in the Hu-PDCoV-specific signature map in the Spike glycoprotein. The first one, P8A, in the N terminal domain of the glycoprotein, is not resolved in the known crystallographic structure, possibly because the segment is too flexible to be seen by cryoEM. The second one, V550A, is located in the S1 subunit (between the RBD and the cleavage site between S1 and S2) on a short beta sheet forming intramolecular contact with a neighboring loop (Fig. 4b). The V550A change observed in Hu-PDCoV (removal of two methyl groups) is present at relatively low frequency in other Asian PDCoVs, neither of which displays the additional amino acid changes observed in the Haitian strains. This change, albeit minor, eliminates specific Van der Waals contact (with Proline at position 535 and the backbone carbonyl at position 532), potentially enhancing protein flexibility and dynamic movement of S1. Since mutations that prevent intermolecular spike protein interactions between S1 and S2 of SARS-CoV-2 variant B.1.1.7 have been observed (A570D, D614G and S982A)^37,38^, V550A may represent a common mechanism that enhances dynamic movements, accelerating virus membrane fusion events and transmission.

**Figure 4.**
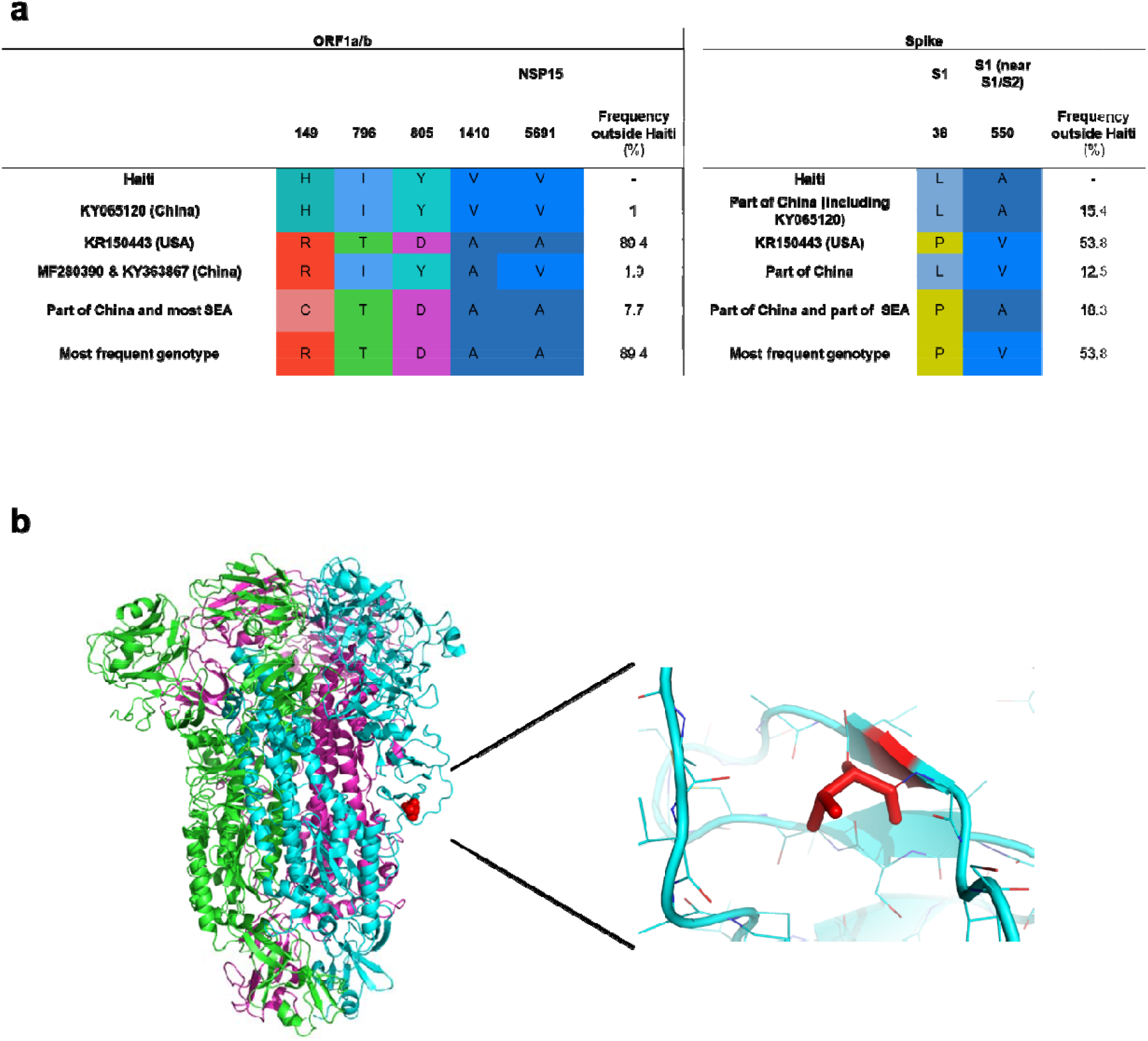
Analysis of conserved amino acids in Hu-PDCoV strains. **a)** Amino acid signature pattern analysis: residues are numbered based on the ORF1a/b absolute amino acid position (starting from number 1) in the reference sequence JQ065043 (Pig/China). The only residues with 100% frequency in Haitian strains mapped in the ORF1a/b and Spike regions. **b)** Spike glycoprotein trimer structure. Different colors are assigned to each monomer. Amino acid residues were numbered relative to their position in the Spike glycoprotein of the reference sequence. Red indicates residue 550, where Haitian sequences have a Valine to Alanine mutation. The other mutation (Proline to Leucine at position 38) was not resolved in the structure.

## Conclusions

To our knowledge, this is the first report of PDCoV infection in humans. The virus was identified in blood plasma samples, consistent with systemic dissemination. While the children in this study did not report diarrhea, two noted abdominal pain, which would be consistent with intestinal involvement; both also reported cough, which may reflect upper respiratory involvement. Since we could not sample from animals in the two regions where the virus was detected, we cannot definitively identify the zoonotic route of the human infections. However, we would note that pigs (often free-roaming) are not uncommon in rural and urban areas of Haiti, and our findings are consistent with a virus maintained in the swine population and capable of successful spillover in humans thanks to the independent emergence of specific amino acid changes through convergent evolution. Further *in vitro* experiments will be necessary for a functional characterization of the Haitian mutants. Yet, the presence of a unique amino acid signature involving both Nsp15 protein and the S glycoprotein, which evolved in parallel along distinct phylogenetic lineages, does suggest a role of these amino acid changes in selection driven adaptation to infection of human hosts.

In pigs, PDCoV is a recently recognized porcine coronavirus which has demonstrated an ability to spread rapidly in animal populations at a global level. Our data highlight the potential for movement of this and other coronavirus strains into human populations, potentially in rural or less-developed regions where contact with domestic animals is common. The children infected with this virus had only mild illness; at this point in time, we do not find evidence that Hu-PDCoV represents a major human health threat. It is of interest, however, that Haiti, despite high levels of infection with SARS-CoV-2^39^, appears to have had less severe COVID-19 diseases than initially expected^40^. Indeed, one hypothesized reason for a decrease in severe SARS-CoV-2 cases in the general population would be the presence of pre-existing immunity to coronavirus strains other than SARS-CoV-2, which may have entered human populations.

## Data Availability

PDCoV sequences are available through GenBank, with GenBank accession numbers provided in Table 1. Data on strains used in the phylogenetic analysis are included in Supplemental Table S1.

## Supplemental Figures and Tables

**Supplemental Table S1** | **Accession numbers of deltacoronaviruses accessed from NCBI**.

**Supplemental Figure S1.**
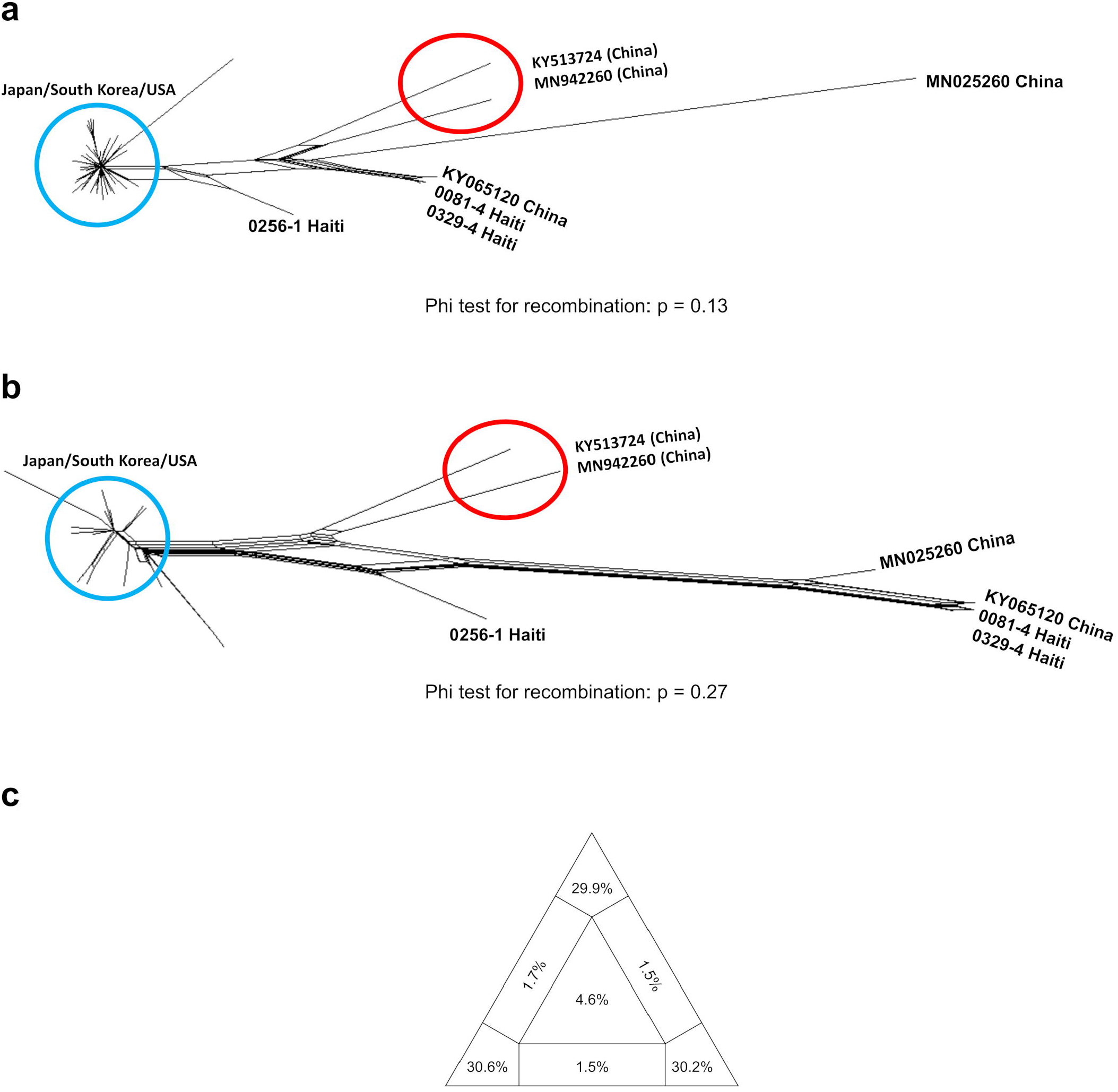
Neighbor net plot of PDCoV fragments and assessment of phylogenetic signal. Neighbor nets were inferred from pair-wise p-distances of 47 genomes for the **a)** major and **b)** minor genome fragment identified by RDP4 (see Methods). Recombination signal was assessed within each fragment by the PHI test. No significant evidence of recombination was found in either fragment (PHI test p > 0.05). The only change in topology, in the split decomposition networks shown in panel a and b, concerned two Chinese sequences (highlighted by the red circle) displaying mixed ancestry possibly due to homoplasy. These sequences were also removed before performing any further phylogenetic analyses. **c)** Likelihood mapping of the final 109 sequences alignment (see Methods) showing extremely low phylogenetic noise (4.6% in the center of the triangle), as required for reliable phylogeny inference.

**Supplemental Figure S2.**
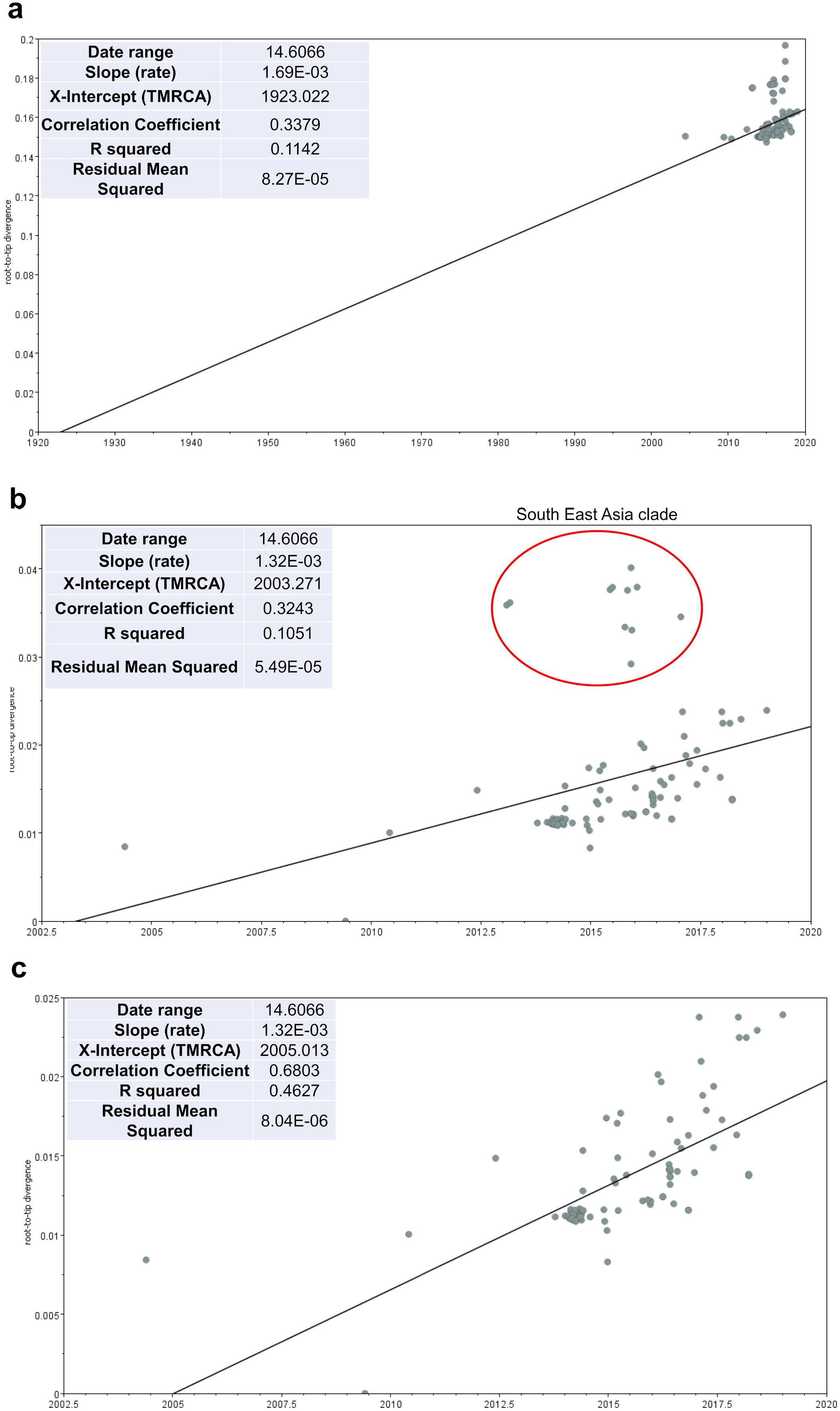
Analysis of the temporal signal with TempEst. Root-to-tip distance (y-axis) *vs*. sampling time linear regression in the ML likelihood phylogeny inferred from: **a)** All PDCoV sequences in the final data set (n = 109) including sparrow outgroup sequences **b)** PDCoV sequences without outgroups (n = 105), with sequences in the red circle belonging to the South East Asia clade (see figure 1) showing a clear departure from the strict molecular clock model, and **c)** PDCoV sequences after removal of the South East Asia clade (n = 94), showing greatly improved clock signal (R^2^ = 0.68).

## Acknowledgments

Work was supported in part by a grant (R01-AI123657S1) from NIAID to JGM.

## Author contributions

JAL supervised virology work, identified the viruses subsequent to performing Sanger sequencing and assembly of the sequences, and contributed to manuscript writing; MST performed genomic and evolutionary analyses and wrote the manuscript; SKW performed virology and molecular work, including GenMark analyses; MAE performed virology and molecular work and coordinated sample shipments and communications between stakeholders in Haiti and the University of Florida; MMA, CJS, TSB, and JCL performed virology and molecular work; TT and SC collected plasma samples, performed medical exams, and documented the findings thereof; CM contributed to bioinformatic analyses and edited the manuscript; VMBDR provided medical oversight over the project and established the internal and external processes including obtaining the necessary permissions for this project; DAO carried out the structural analysis; JAL, MS and GM designed the study and contributed to manuscript writing.

## Data availability statement

GenBank accession numbers for sequence data are included in Table 1. Supplementary Table S1 includes a listing of accession numbers of deltacoronaviruses accessed from NCBI for the phylogenetic studies.

## Methods

### Clinical Sample Collection

From 2012 to 2020, our research group monitored a cohort of approximately 1250 school children in the Gressier region of Haiti^21^. Children attended one of four schools operated by the Christianville Foundation and had free access to medical care through a school-based clinic.

Children presenting to the clinic with an acute undifferentiated febrile illness, defined as a history of fever and/or a measured temperature > over 37.5°C in the clinic with no localizing symptoms or signs (i.e., no respiratory, skin, or urinary symptoms or signs) were invited to enroll in an ongoing study of acute febrile illness; written informed consent for study participation was obtained from parents, with assent from children. After enrollment, clinic health care providers recorded clinical data in a study questionnaire and a sample of venous blood (1-3 ml) was collected in an acid-citrate-dextrose blood collection tube. The blood samples were subsequently centrifuged to pellet the platelets, red blood cells, and white blood cells, and the resulting plasma was aseptically transferred to cryovials and stored at −80°C for subsequent analysis. Appropriate medical care based on clinical presentation and laboratory studies was provided to study participants by clinic health care providers. Data on identification of arboviruses and other virus species among children participating in the study have been previously reported^22,23,25,26,41-44^.

Studies were approved by the Institutional Review Board (IRB) at the University of Florida and the Haitian National IRB. As the study was done in young children the amount of plasma collected was limited, and samples have, in most instances, been exhausted, due to the range of studies initially conducted on the samples while screening for other pathogens. IRB restrictions limit our ability to share samples outside of our institution.

### Virus Identification and Sequencing

Attempts at next-generation sequencing using an Illumina MiSeq platform generated minimal coverage, so we sorted to Sanger Sequencing using the primer system outlined by Liang et al^45^, with one addition: to obtain the 5′ ends of the viral genomes, a Rapid Amplification of cDNA Ends (RACE) kit was used per the manufacturer’s protocols (Life Technologies, Carlsbad, CA, USA), and the resulting amplicons TA-cloned into plasmids and sequenced. PCR amplicons for Sanger Sequencing were amplified using AccuScript High-Fidelity reverse transcriptase (Agilent Technologies, Inc., Santa Clara, CA) in the presence of SUPERase-In RNase inhibitor (Ambion, Austin, TX), followed by PCR with Q5 DNA polymerase (New England Biolabs. They were next purified using a QIAquick PCR purification kit (Qiagen Inc., Germantown, MD) before TA-cloning. The inserts in the plasmids were subsequently sequenced bidirectionally using a gene-walking approach, based on obtaining at least 800 bp or non-ambiguous sequence. Briefly, pairs of non-overlapping primers and Q5 polymerase were used to produce 42 separate amplicons corresponding to the PDCoV genome, and each amplicon was Sanger sequenced bidirectionally.

### Sequence data assembly

The identity of the whole genome sequences was confirmed *via* online BLAST^46^ of the nr/nt NCBI database. Following positive identification, available PDCoV sequences from pigs were downloaded from NCBI (www.ncbi.nlm.nih.gov), together with closely related sparrow deltacoronavirus sequences^47^ to be used as outgroups in the phylogenetic analysis (see below). The final full genomes data set assembled included 104 PDCoV genomes from pigs, 4 from sparrows (Supplemental Table S1), and 3 newly sequenced Hu-PDCoV strains, which were aligned with MAFFT^48^v.7.407. Potential recombinant origin of Haitian sequences was assessed using an algorithm based on the PHI test^49,50^ implemented SplitsTree4^30^ v.4.14.8 and with the RDP4^31^ software package v.4.97. RDP4 detected several possible recombination events in the full data set (although homoplasy could not be ruled out), resulting in at least two major breakpoints at nucleotide position 14,874 – 17,234 (numbered according to reference sequence JQ065043) that involved 60 strains from Chinese pigs belonging to sequence clusters unrelated to the new human isolates. After such strains were removed from the alignment, together with the outgroup sequences, the PHI test on the remaining subset of 47 sequences (44 from pigs + 3 new human strains) still detected evidence of recombination (p<0.001). Split decomposition analysis using SplitsTree5 of the major (1 – 14873/17235 – 25,413) and minor recombinant fragment (14,874 – 17,234) identified two additional Chinese sequences (KY513724 and MN942260) with mixed ancestry, possibly due to homoplasy, that were removed from the full data set. However, since the other Chinese pig strains of possible recombinant origin clustered in clades clearly distinct from the ones including the human isolates, and our goal was to assess the most recent common ancestor between human and most closely related pig strains, a final alignment (available from the authors upon request) including 109 sequences (4 outgroup sequences from sparrow, 102 from pigs and 3 from human patients) was chosen to carry out subsequent phylogenetic analyses.

### Phylogenetic and amino acid signature analysis

Phylogenetic signal was verified using likelihood mapping^51^, as implemented in IQTREE v.2.0.6^52^. Maximum likelihood tree was calculated using the same version of IQTREE, with the best fitting nucleotide substitution model according to the Bayesian Information Criterion (BIC) and 1,000 bootstrap replicates. The correlation between root-to-tip genetic divergence and sampling dates to assess clock signal of the alignment were performed with TempEst^32^ prior to Bayesian phylodynamic analyses. The time-scaled tree was calculated using the Bayesian phylodynamic inference framework in BEAST v. 1.10.4^53^;

Markov chain Monte Carlo (MCMC) samplers were run for 200 million generations, with sampling every 20,000 generations, to ensure proper mixing, which was assessed by calculating the effective sampling size (ESS) of each parameter estimate. The HKY nucleotide substitution model was used with empirical base frequencies and gamma distributions of site-specific rate heterogeneity^54^. The molecular clock was calibrated by enforcing a strict clock and and chosing either a constant size or a Bayesian Skyline Plot demographic prior^55^. A maximum clade credibility tree was inferred from the posterior distribution of trees using TreeAnnotator, specifying a burn-in of 104 million and median node heights. The MCC was edited graphically using *ggtree* ^56-58^. MCMC runs with different demographic priors gave the same result.

Signature pattern analysis of the Haitian strains as compared to the rest of the downloaded sequences was performed with online version of VESPA^33^; the PDCoV reference sequence JQ065043.2 was used as a guide for the codon coordinates. The spike glycoprotein three-dimensional structure PDB 6b7n^59^ was used as a base, and figures were generated using PyMol^60^.

